# Knowledge representation of a multi-centre adolescent and young adult (AYA) cancer infrastructure; development of the STRONG AYA Knowledge Graph

**DOI:** 10.1101/2025.06.03.25328788

**Authors:** J. (Joshi) Hogenboom, V. (Varsha) Gouthamchand, C. (Charlotte) Cairns, S.H.M. (Silvie) Janssen, K. (Kirsty) Way, A.L.A.J. (Andre) Dekker, W.T.A. (Winette) Van Der Graaf, A. (Anne-Sophie) Darlington, O. (Olga) Husson, L.Y.L. (Leonard) Wee, J. (Johan) Van Soest, A. (Aiara) Lobo Gomes

## Abstract

**Purpose:** Rare diseases are difficult to fully capture, and regularly call for large, geographically dispersed initiatives. Such initiatives are often met with data harmonisation challenges. These challenges render data incompatible and impede successful realisation. The STRONG AYA project is such an initiative, specifically focusing on adolescents and young adults (AYAs) with cancer. STRONG AYA is setting up a federated data infrastructure containing data of varying format. Here, we elaborate on how we used healthcare-agnostic Semantic Web technologies to overcome such challenges.

**Methodology:** We structured the STRONG AYA case-mix and core outcome measures concepts and their properties as knowledge graphs. Having identified the corresponding standard terminologies, we developed a semantic map based on the knowledge graphs and the here introduced annotation helper plugin for Flyover. Flyover is a tool that converts structured data into Resource Descriptor Framework (RDF) triples and enables semantic interoperability. As a demonstration, we mapped data that is to be included in the STRONG AYA infrastructure.

**Results:** The knowledge graphs provided a comprehensive overview of the large number of STRONG AYA concepts. The semantic terminology mapping and annotation helper allowed us to query data with incomprehensible terminologies, without changing them. Both the knowledge graphs and semantic map were made available on a Hugo webpage for increased transparency and understanding.

**Discussion:** The use of Semantic Web technologies such as RDF and knowledge graphs are a viable solution to overcome challenges regarding data interoperability and reusability for a federated AYA cancer data infrastructure without being bound to rigid standardised schemas. The linkage of semantically meaningful concepts to otherwise incomprehensible data elements demonstrates how by using these domain-agnostic technologies we made non-standardised healthcare data interoperable.

## Introduction

Adolescent and young adult (AYA) cancer is rare and concerns an often-overlooked population, defined as people aged 15 to 39 years at primary cancer diagnosis. AYAs are in part characterised by significant differences in tumour type, psychosocial characteristics and care needs. As a result, AYAs cancer calls for age-specific care that is traditionally unmet by paediatric and adult cancer care^1^.

To improve healthcare services, research, and outcomes for AYAs, the STRONG AYA Initiative^2^ is setting up a federated data infrastructure^3,4^ that incorporates both retrospective and prospective AYA data – contributed by several medical centres across Europe. This regional variability allows us to highlight significant challenges in data harmonisation across healthcare systems.

The definitions, format, and terminology of the data in these datasets vary across institutions; this is often for practical and operational purposes. For example, when recording an individual’s highest obtained educational level, it is most pragmatic to consult participants in a format that is sensible in their regional setting – as this is the information people will know. Such differences can render data incompatible with that collected in other regional settings, if not adequately harmonised.

Therefore, one of the first crucial steps in such large and international collaborations is to adopt or establish certain standards within a data model. This involves two distinct components: standardising the data schema, and standardising the definitions and terminology. Establishing a standardised schema provides syntactic interoperability; but this still relies on semantic interoperability – standardising definitions and terminology. These two components constitute interoperability in the broader sense, enabling data integration from diverse sources^5^. For educational level, a straightforward solution would involve transcribing such regionally sensible levels to UNESCO’s International Standard Classification of Education (ISCED) ^6^, but this can be costly and burdensome. Moreover, even for simple concepts, intrinsic differences in data semantics regularly occur. For example, *biological sex* can be recorded as *‘male, female’*, *‘male, female, intersex’*, and *‘0, 1, 2’*. More complex cases are abundant, and even a seemingly straightforward example like the *time of diagnosis* can quickly reveal operational differences. The definition of the *time of diagnosis* can range from the first illness-related hospital visit to the date of biopsy evaluation or formal diagnosis made by a clinician.

A well-established approach to solve such challenges is the implementation of the F.A.I.R. – findable, accessible, interoperable, and reusable – data guiding principles^7^. This previous work defines how data can be made F.A.I.R. for both humans and machines, whilst allowing flexibility in terms of multiple co-existing semantic ontologies schema and semantics. In a federated data infrastructure, applying the F.A.I.R. principles has been effective in overcoming interoperability hurdles related to data semantics^8,9^, while bridging pitfalls concerning syntactic interoperability e.g. using an ‘on-read’ approach^9^. At the same time, the F.A.I.R. principles highlight reusability, thus increasing the understanding of data – and hence transforming it into information – which is a pivotal aspect of the process. To that end, the use of knowledge graphs and Semantic Web standards are established methods as they are semantically rich and reflect the structure of the data at hand^8–12^. These concepts represent complex information in a graphical format and, therewith, aim to enhance understanding of the data by illustrating relationships between data concepts. These methodologies are however domain agnostic and lack the specificity relevant for STRONG AYA.

The aim of this work is to develop and implement a tailored data model for STRONG AYA that addresses the unique challenges of AYA cancer data harmonisation. To achieve this, we developed a data model for STRONG AYA that is aligned with its data collection procedures, simultaneously delivering on the implementation of the ’Interoperable’ and ’Reusable’ aspects of the F.A.I.R. data principles. In this work, we elaborate on this effort to illuminate the various processes involved with resolving data incompatibilities in a large healthcare consortium’s federated infrastructure. To reduce the complexity of concepts relevant for AYAs with cancer, we developed the STRONG AYA data model as a knowledge graph. Using this knowledge graph, we then transcribe its contents to the STRONG AYA semantic map which can be used for the necessary mapping that ensures semantic interoperability, whilst overcoming syntactic interoperability through an established ‘on-read’ approach. With the interplay of the knowledge graph and semantic map we aim to accelerate and facilitate STRONG AYA’s goals of improving healthcare services, research, and outcomes for AYAs – whilst also setting an example of F.A.I.R. data principles implementation in large scale consortia.

## Methodology

### Data elements

In STRONG AYA, extensive research identified key information to enhance healthcare services, research, and outcomes for AYAs with cancer. This involved a literature review, qualitative interviews, and a three-round Delphi procedure with AYA cancer stakeholders (AYAs with cancer, caregivers, health professionals, researchers, and policymakers) to determine relevant outcome domains^13,14^. A Core Outcome Set (COS) was developed from these domains. Subsequently, a set of core measurement instruments and/or items were compiled which best measure the COS. The COS and measurement set were refined to minimise participant burden while retaining essential elements. A list of relevant case-mix variables, identified through a literature review^15^, supplemented the COS to form the final data elements for the STRONG AYA infrastructure, represented in table 1, excluding all time elements except the initial timestamp. Details on these procedures can be found in their original publications^13–15^.

**Table 1:**
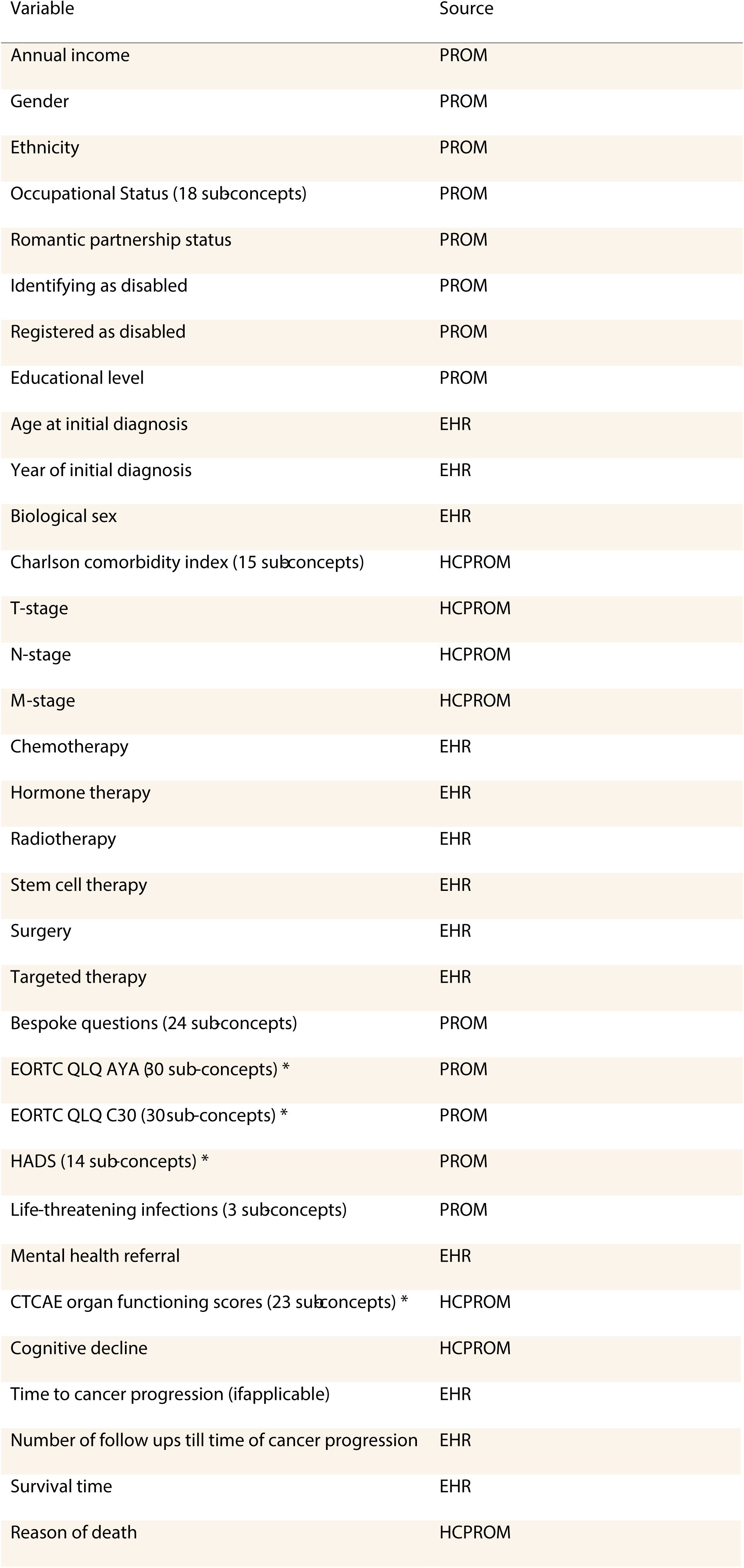
Overview of AYA cancer relevant concepts compiled through a literature review, qualitative interviews, and a three-round Delphi procedure with AYA cancer stakeholders.

| Variable | Source |
| --- | --- |
| Annual income | PROM |
| Gender | PROM |
| Ethnicity | PROM |
| Occupational Status (18 subconcepts) | PROM |
| Romantic partnership status | PROM |
| Identifying as disabled | PROM |
| Registered as disabled | PROM |
| Educational level | PROM |
| Age at initial diagnosis | EHR |
| Year of initial diagnosis | EHR |
| Biological sex | EHR |
| Charlson comorbidity index (15 subconcepts) | HCPROM |
| T-stage | HCPROM |
| N-stage | HCPROM |
| M-stage | HCPROM |
| Chemotherapy | EHR |
| Hormone therapy | EHR |
| Radiotherapy | EHR |
| Stem cell therapy | EHR |
| Surgery | EHR |
| Targeted therapy | EHR |
| Bespoke questions (24 subconcepts) | PROM |
| EORTC QLQ AYA β0 sub-concepts) * | PROM |
| EORTC QLQ C30 (30sub-concepts) * | PROM |
| HADS (14 sub-concepts) * | PROM |
| Life-threatening infections (3 subconcepts) | PROM |
| Mental health referral | EHR |
| CTCAE organ functioning scores (23 subconcepts) * | HCPROM |
| Cognitive decline | HCPROM |
| Time to cancer progression (ifapplicable) | EHR |
| Number of follow ups till time of cancer progression | EHR |
| Survival time | EHR |
| Reason of death | HCPROM |

### Data conversion and annotation

For multi-centre semantic mapping and knowledge graph generation we used the Resource Description Framework (RDF) data format^16^. RDF is a data representation standard for the basic building block of a graph: the representation of nodes and arcs. This triple format is made up of a subject – predicate – object statement representing node – arc –node, respectively. For instance, *‘AYA – has column – biological sex’*.

As none of the centres collected data as RDF-triples, we used the *Flyover* tool^9^; (https://github.com/MaastrichtU-CDS/Flyover) to harmonise the data format across centres. *Flyover* converts an arbitrary form of data such as comma separated values (CSV), into triples and then stores them in a graph database. For instance, a row – AYA 1 – with a value of ‘*femalè* for *‘biological_sex’* being converted into *‘AYA 1 – has column – biological_sex’* and *‘biological_sex – has value – femalè* as is illustrated in figure 1.

**Figure 1:**
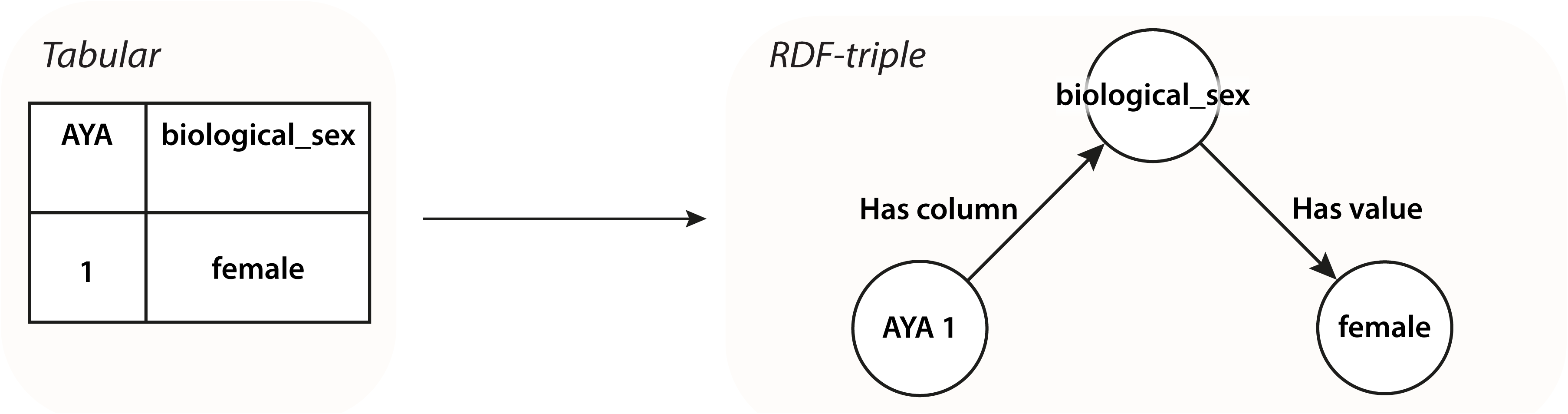
Conversion of tabular biological sex data to RDF-triple format using Flyover.

Using this triple format, *Flyover* allows us to impose semantics on top of this existing data through a metadata layer – or annotation graph. This means that the *‘AYA – has column – biological_sex’* triple can refer to variables whose names do not necessarily carry semantic significance, which is one of the challenges that was emphasised in the introduction. *Flyover* maintains the original data structure by providing semantic interoperability on-read through this annotation graph.

To make the best use of *Flyover*’s descriptives abilities, we developed a *JSON* semantic map plugin for *Flyover*’s graphical user interface. Using this semantic map, we can directly map variable names as they appear in their original data source, to standardised terminologies. This semantic map could then be used to easily develop the queries that annotate the original data sources’ names in the metadata layer through *Flyover*’s *Annotation Helper*. This helper parses the variable and values names’ along with the mapped standard terms into queries that insert triple statements into the annotation graph.

### Knowledge graphs and semantic map

The RDF model enabled the addition of semantically rich graph structures to the data without modification. Using the list of AYA cancer-relevant concepts, we created a visual knowledge graph to illustrate these data elements in a structured way. We displayed this visual knowledge graph in three sub-structures, all part of a single graph model: data, data source, and instrument graph. The graph structure was then reviewed by those who defined the list of AYA cancer relevant concepts elements. After this review, we included the graph structures in the semantic map so that they could be incorporated in the metadata layer.

For the STRONG AYA data elements, we identified relevant standardised terminologies, predominantly leveraging the *National Cancer Institute Thesaurus* (NCIt) ^17^ for object terms – or classes, and the *Semanticscience Integrated Ontology* (SIO) ^18^ for properties. Other vocabularies that were used include the *Gender, Sex, and Sexual Orientation Ontology* (GSSO) ^19^ and *SNOMED CT* (SCT) ^20^. We used custom terms for STRONG AYA’s bespoke questions and concepts lacking standardised definitions. We used *Flyover*’s *Annotation Helper* semantic map format as basis and as overview of all data elements and their standard terms. This global semantic map – without local terms – was published on GitHub to allow for transparent semantic map updates and traceability. The workflow that was used to achieve semantic interoperability is illustrated in figure 2.

**Figure 2:**
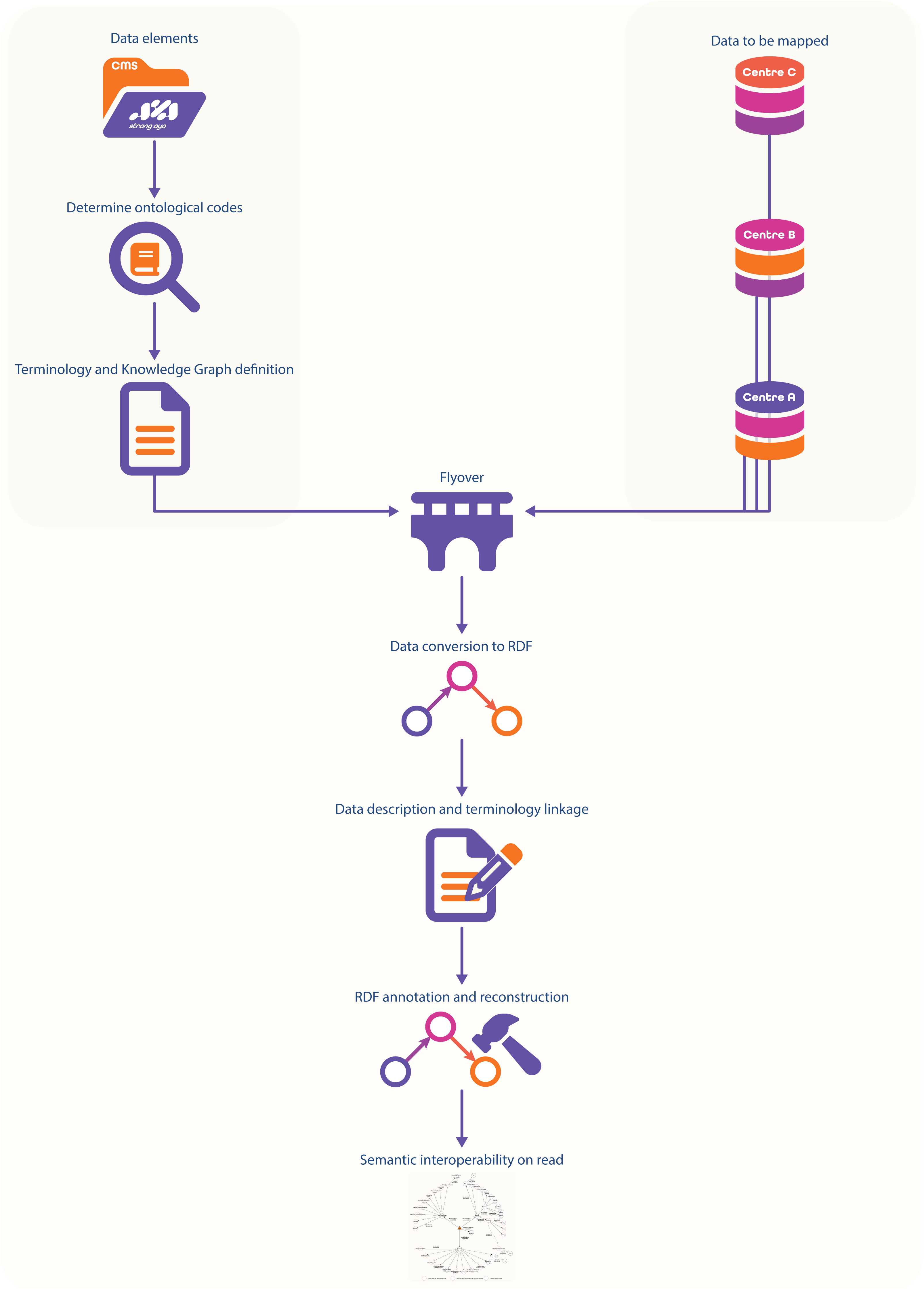
Workflow used to achieve semantic interoperability for STRONG AYA data elements and data-contributing centres.

To provide a comprehensive overview of our knowledge graph and semantic map we integrated them into a STRONG AYA Knowledge Representation, semi-static *Hugo* (https://gohugo.io) website. This resource enables continuous review by consortium members. The semantic map section displays variable names, vocabulary reference codes, and associated preferred names and definitions from BioPortal (https://bioportal.bioontology.org). Content is updated quarterly and upon GitHub repository update by extracting semantic map information and fetching vocabulary details via the BioPortal REST API. Unfound reference codes are automatically reported as GitHub issues to alert repository owners.

### Semantic mapping demonstration

As part of STRONG AYA’s retrospective data retrieval and to demonstrate our semantic mapping method, we mapped the SURVAYA study (ClinicalTrials.gov identifier: NCT05379387), a population-based cross-sectional cohort study of long-term AYA cancer survivors from the Netherlands Cancer Registry. Details can be found in the original study publication^21^. SURVAYA will be integrated into STRONG AYA’s infrastructure, but for testing, we used a synthetic dataset^22^ containing overlapping elements. The SURVAYA study was conducted in accordance with the Declaration of Helsinki and approved by the Netherlands Cancer Institute Institutional Review Board (IRBIRBd18122) on February 6, 2019. The synthetic dataset was used with permission from the study’s principal investigator and sponsor.

## Results

### Knowledge graphs

The knowledge graph in figure 3 offers a visual and structured representation of AYA cancer-relevant concepts. Specifically, figure 3 presents the data graph, categorising data concepts into sociodemographic, clinical, and outcome characteristics using – using SIO’s ‘has annotation’ – to reduce complexity and structure concepts. Data concepts are generally attributes of a given category, utilising SIO’s ‘has attribute’. Units for continuous concepts, such as age at diagnosis, overall progression time, and survival, were added in years and days to enhance interpretability, associated via SIO’s ‘has unit’. Intervariable relationships are limited to neoplasm-associated concepts, including cancer progression time, tumour staging, and localisation, – which utilise SIO’s ‘is related to’ and ‘has property’. The AYA’s research identifier is directly associated with the AYA using SIO’s ‘has unique identifier’ and is not part of any sub-category. Data sources are colour-coded: orange for patient-reported outcome measures (PROMs), pink for healthcare professional reported outcome measures (HCPROMs), and purple for electronic health records (EHRs).

**Figure 3:**
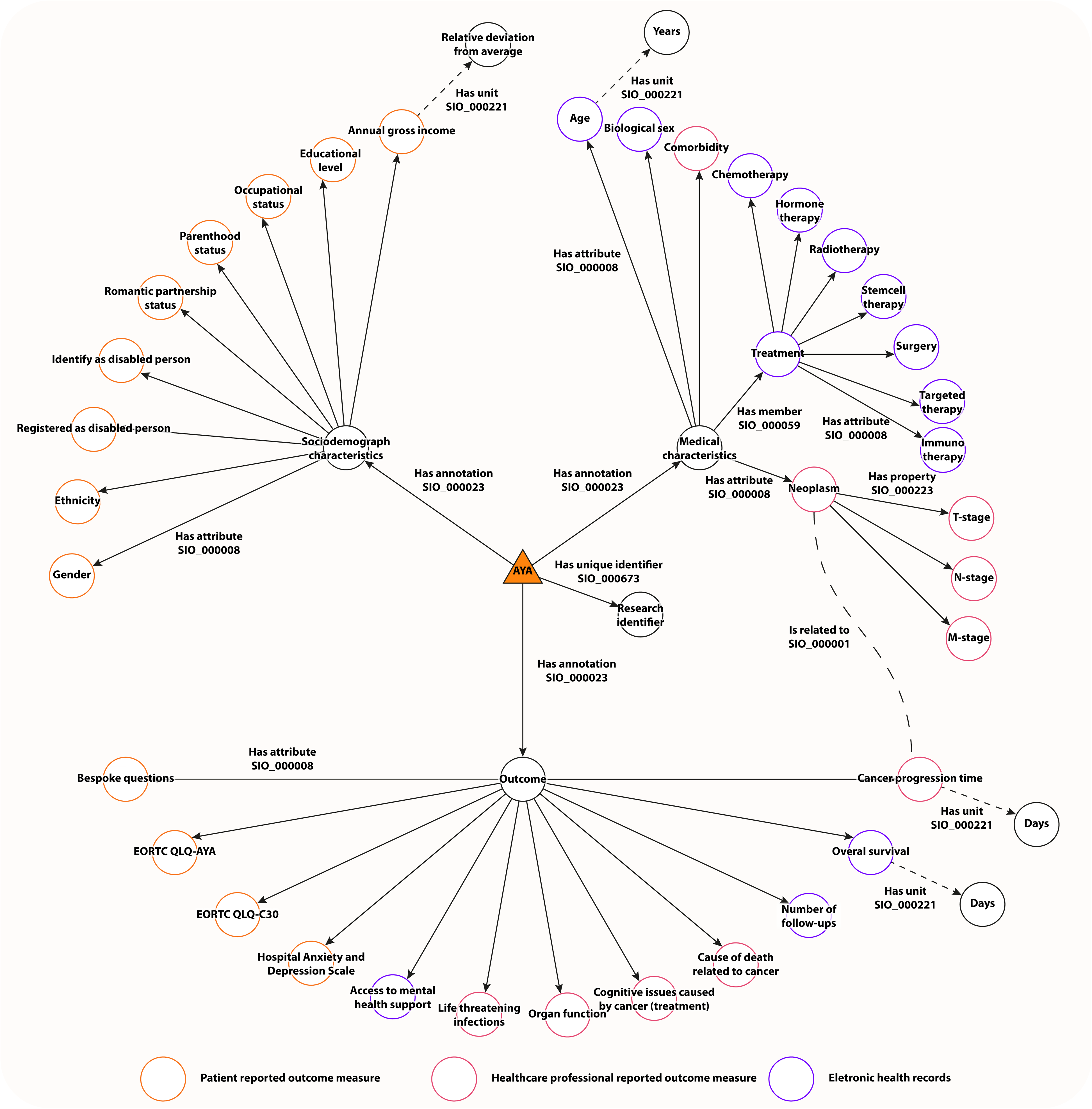
Data graph showcasing the AYA cancer relevant concepts in a more comprehensive way. Information on collection time is not present here and is visible in the underlying instrument graphs. The measurement instrument type or data source per concept is identifiable by the coloured outline. Please note that while certain concepts are specifically categorised as ’Outcome,’ what constitutes an outcome is study-specific and may also include variables categorised here under ’Medical characteristics’ and ’Sociodemographic characteristics.

The data source graphs are displayed in supplementary figure 1 and describe the data elements’ sources using SIO’s ‘has property’, detailing the distinct properties of PROM, HCPROM, and EHR data elements. Supplementary figure 2 introduces an additional layer to the graph structure by clustering data related to a single concept, exemplified through the European Organisation for Research and Treatment of Cancer Quality of Life Questionnaire for AYAs and its specific questions^23^.

### Knowledge representation webpage

Figure 4 shows an excerpt of the semantic map and the knowledge representation webpage of the concept *biological sex*. The semantic map describes the references to standard vocabularies (in bold orange font) and defines the graph structure (in bold blue font). Concretely, this semantic mapping annotates our triple statement of *‘AYA 1 – has column – biological_sex’* with *‘AYA 1 – sio:SIO_000235 – ncit:C18772’* and *‘ncit:C18772 – sio:SIO_000008 – ncit:C28421’*.

**Figure 4:**
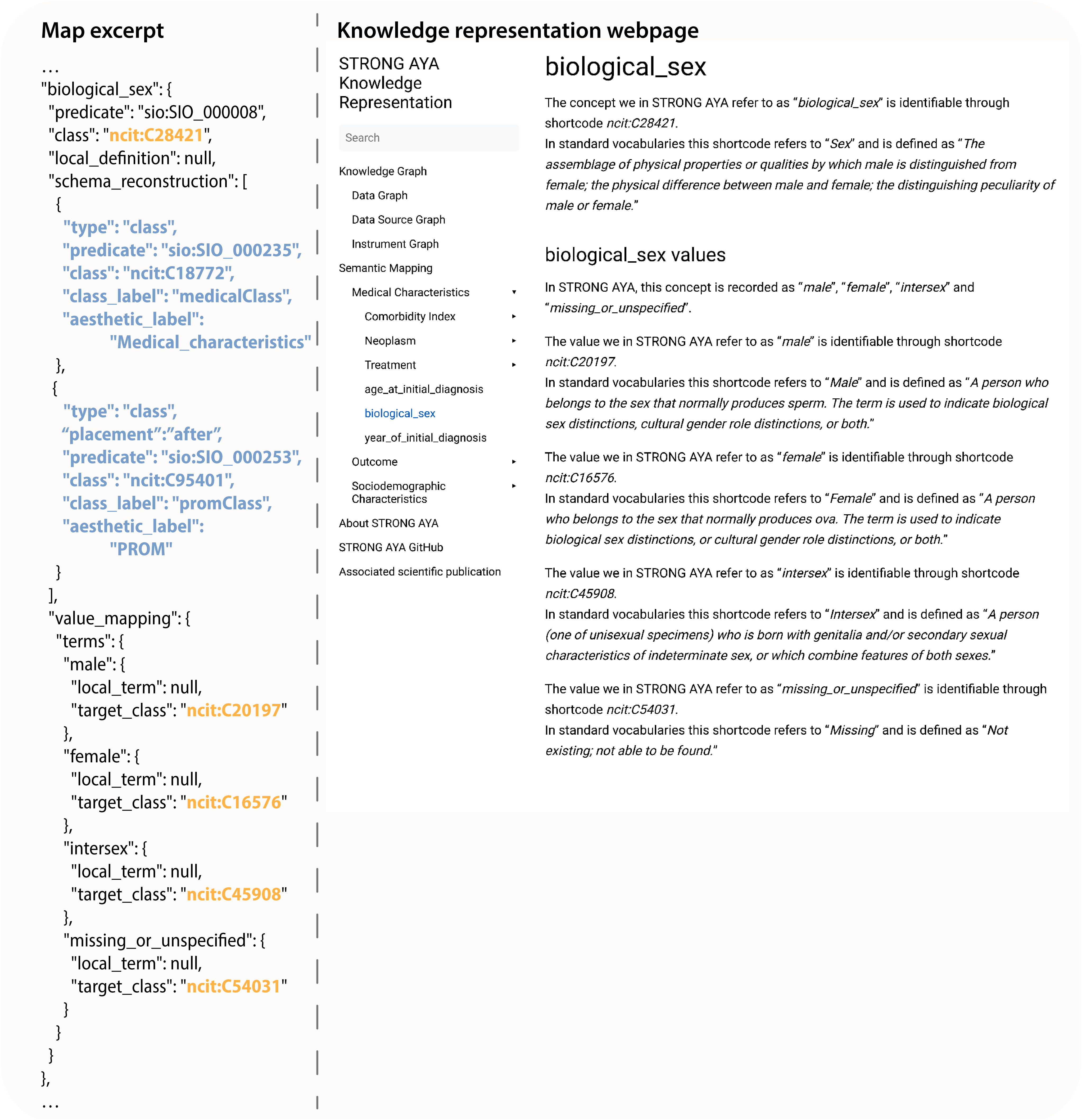
An excerpt of the AYA semantic map and of the AYA cancer knowledge representation. With in the semantic map excerpt the reference to standardised terminologies in bold orange font,

The single triple statement is reconstructed to two statements through the schema reconstruction section, which reflects the structure of the previously described knowledge graphs. Our value triple statement of *‘biological_sex – has value – femalè* is annotated with *‘ncit:C28421 – has value – ncit:C16576’.* Whilst in the webpage the structure is displayed in a human-readable format by showing what the references in the semantic map – here being the triple statements classes – correspond to.

The complete semantic map (https://github.com/STRONGAYA/AYA-cancer-semantic-map) and the knowledge representation pages (https://strongaya.github.io/AYA-cancer-semantic-map) are available on GitHub.

### Semantic mapping demonstration

The interoperability of the annotated RDF-triple SURVAYA data is illustrated in figure 5, showcasing data accessibility through both local and standard terminology – here exemplified using biological sex. Utilising Flyover and the semantic map, we tested our data harmonisation workflow with synthetic SURVAYA data. Initially, RDF-converted biological sex data of a SURVAYA dataset would solely be available as *‘AYA 1 – has column – alg_v1b’*, but through annotation this data becomes accessible through the standard semantic mapping of *‘AYA 1 – sio:SIO_000235 – ncit: C326200’* and *‘ncit:C326200 – sio:SIO_000008 – ncit:C28421’*. SPARQL^24^ was used for data queries. The semantic mapping for SURVAYA is available on GitHub (https://github.com/STRONGAYA/AYA-cancer-semantic-map/tree/dev/retrospective/SURVAYA). Supplementary figure 3 presents the semantic mapping excerpt for biological sex data, emphasising the necessity of semantic meaning, as local terminology lacks clarity without it. The full output of our graph database containing synthetic SURVAYA data is available in supplementary figure 4.

**Figure 5:**
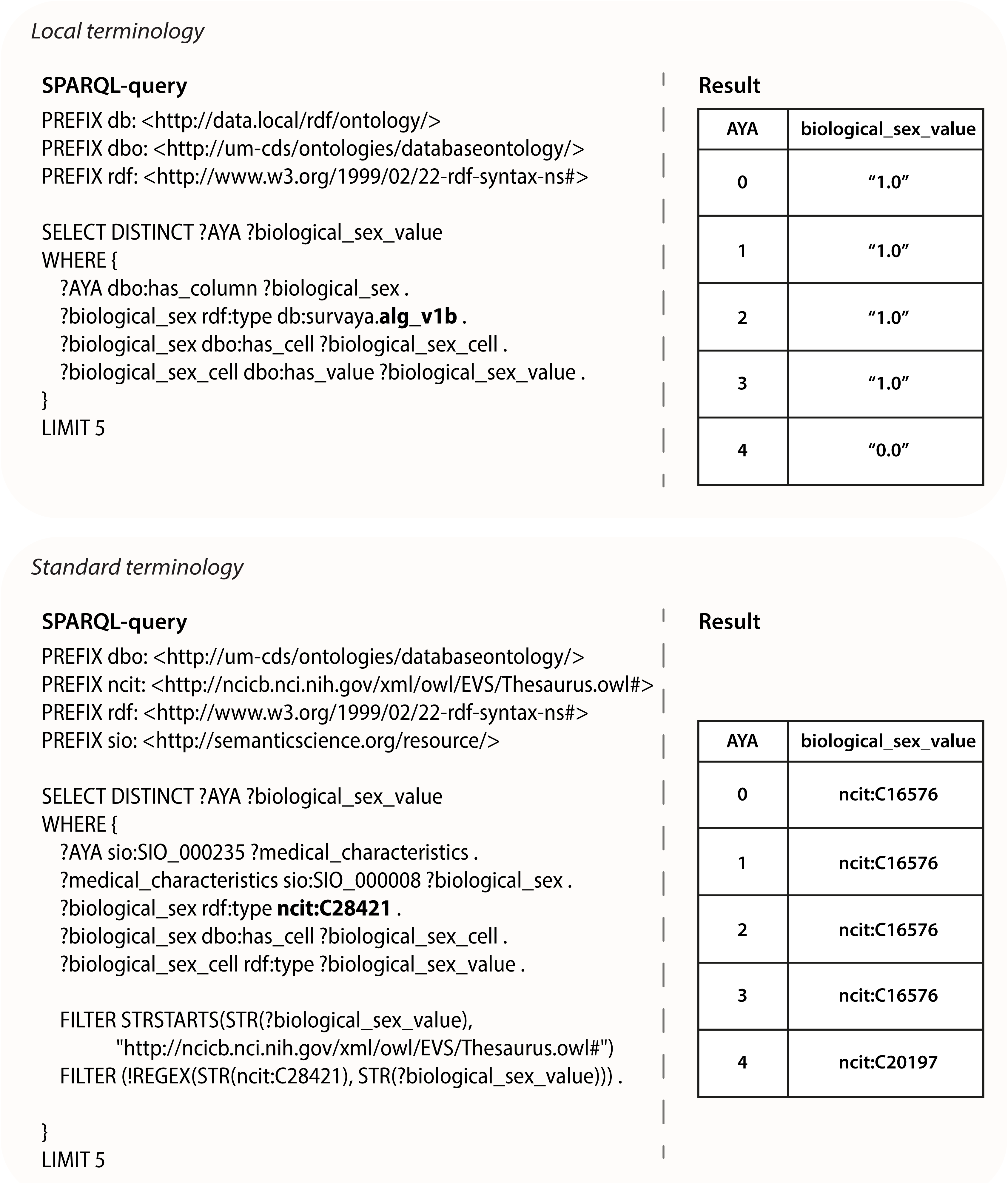
Demonstration of the interoperability of the SURVAYA dataset, which through a SPARQL-query is both accessible via its original – and incomprehensible– terminology ’alg_v1b’ and standardised terminology ’ncit:C28421’. Both the local terminology and the standard terminology are highlighted in bold font. Shown values are from synthetic SURVAYA data and do not contain information of real participants.

## Discussion

The knowledge representation described in this work illustrates how, by making use of RDF-data structures, we can make a large mix of complex, AYA cancer concepts to adhere to the ’Interoperable’ and ’Reusable’ aspects of the F.A.I.R. data principles^7^. Our work demonstrates that adhering to these principles allows us to navigate through the difficulties of heterogeneous semantics and inherent differences in data schemas whilst simultaneously expanding the application of a domain-agnostic standard such as RDF.

The STRONG AYA Knowledge Graph has provided a comprehensive overview of the large number of items collected for this consortium. This knowledge graph was then transcribed into a STRONG AYA data semantic map. In turn, the introduction of this semantic map enables us to map and annotate AYA-specific concepts with standardised terminologies, thereby circumventing the use of project-specific definitions. We showcase how we use the AYA knowledge representation on one of the AYA datasets to be included in STRONG AYA, laying a foundation for other datasets.

In the process of making data more F.A.I.R., there are numerous approaches to address challenges related to data interoperability and enhance understandability. In this work, we used Semantic Web standards, such as RDF^16^ and SPARQL^24^, due to the flexible and non-rigid schema design of the RDF format. This flexibility allows data-contributing partners to submit data in its original format, with most interoperability work managed by a single coordinating party. Partners can use the Flyover tool and Annotation Helper plugin with the STRONG AYA semantic map for most scenarios. The introduced Annotation Helper significantly simplifies this process, providing an easy-to-understand aid verifiable by those without RDF or Semantic Web knowledge. The annotation helper also enhances RDF’s flexibility, as the annotation layer can be reapplied as requirements evolve. For instance, the introduced instrument graph supports simultaneous use of cross-sectional and longitudinal data, but as prospective data collection procedures evolve, the instrument graph must adapt. Additionally, the RDF format allows for future inclusion of logical reasoning to identify erroneous combinations in the STRONG AYA Knowledge Graph, benefiting data quality assurance and efficient data use.

In comparison to other data models, such as the Observational Medical Outcomes Partnership Common Data Model (OMOP-CDM) ^25^, our approach benefits from *Flyover*’s ‘on-read’ approach^9^, allowing the STRONG AYA knowledge graph and schema to adapt without modifying the data. Additionally, inherently to Semantic Web standards and the main mantra “Anyone can say Anything about Anything” we are not limiting ourselves to given terminology standards and data schemas. By using uniform resource identifiers (URIs) of terms – which should resolve to their descriptions – these standards are more open to extension by anyone. However, this flexibility means forgoing the tools available for OMOP-CDM, which, despite its rigidity, offers a well-established ecosystem for data integration and analysis. In contrast, RDF provides the flexibility crucial for STRONG AYA’s diverse and evolving data structures. However, rather than comparing RDF to OMOP, they should be considered complementary. RDF is a data representation standard predominantly used by big-tech and industry^26,27^, while OMOP is a healthcare-specific data model. Future work should focus on integrating OMOP and RDF to create a sustainable hybrid solution.

When developing any form of knowledge representation, it is vital that the included concepts are relevant to the overarching subject. In our work, we have based our AYA cancer knowledge representation on an extensive Delphi procedure and literature review^13,15^. This procedure significantly reduced difficulties in defining the relevant concepts – reiterating the importance of such preparatory work. Adherence to this protocol ensured that the true meaning of a term such as *‘date of diagnosis’* was already quite refined, and prevented extensive discussions during data model development. Because of this robustness and rigorous adherence to these predefined concepts, we were required to use numerous custom, and thus non-standard terminologies. This is because a substantial number of AYA cancer concepts are not present in any ontology, owing to the bespoke nature of various PROMs. Although this does not hinder interoperability, the lack of standard terms diminishes understandability, as our custom ontology codes have no established semantic significance. Moreover, while this work advances knowledge representation for AYA cancers, it also highlights areas lacking common definitions. It underscores both the need for extended AYA research and the establishment – and adoption – of standard PROM terminologies^28^ .

All in in all, we have shown how developing an AYA knowledge representation can navigate the challenging topography of interoperability and understandability in a large AYA cancer consortium. By leveraging existing tools and terminologies, we can increase the adherence of our AYA data pool to the F.A.I.R. data principles whilst concurrently reducing the burden for our data-contributing partners by using *Flyover*’s integrated *Annotation Helper*.

Whilst issues of incompatibility and understandability seem addressed, further implementation of F.A.I.R. data principles will enhance the societal benefits of the data collected in STRONG AYA. Future work should focus on transcribing our knowledge representation to a machine-findable source that, through appropriate agreements, licenses, and protocols is accessible to individuals currently outside of the consortium – simultaneously addressing the ‘Findable’ and ‘Accessible’ attributes in F.A.I.R. To maximise the reusability for a wider audience it is however a necessity that future work also focuses on the introduction of standardised terminology for AYA cancer – and other fields with notable reliance on PROMs.

## Data Availability

The knowledge graph, semantic map, and knowledge representation website are available on GitHub: https://github.com/STRONGAYA/AYA-cancer-semantic-map.

## Acknowledgements

All individuals who have contributed to this study are included in the author list.

