## Supplementary Figure 1 for "Knowledge representation of a multi-centre adolescent and young adult (AYA) cancer infrastructure; development of the STRONG AYA Knowledge Graph"

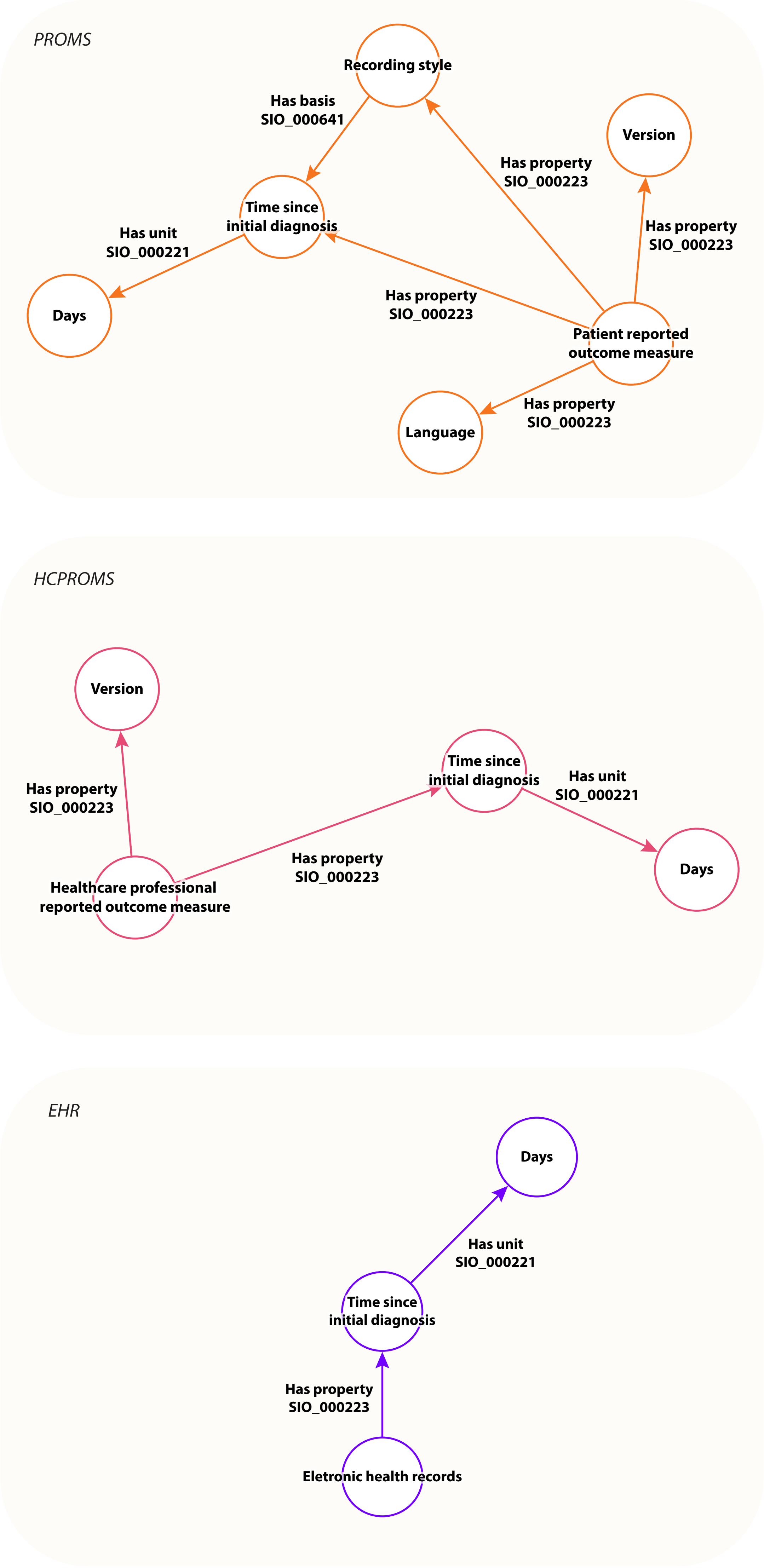


Supplementary Figure 1: Instrument graphs showcasing the underlying structures of the various measurement instruments – or data sources – in the AYA cancer knowledge representation. Each source type had distinct properties, but all included a variable for time elapsed since diagnosis, derived from the recording timestamp, with a unit of days via SIO's 'has unit' property. PROMs and HCPROMs included a 'version' property, with PROMs also featuring a 'language' property. Each data source was linked to its data element using SIO's 'has source' property.
