## Supplementary Figure 2 for "Knowledge representation of a multi-centre adolescent and young adult (AYA) cancer infrastructure; development of the STRONG AYA Knowledge Graph"

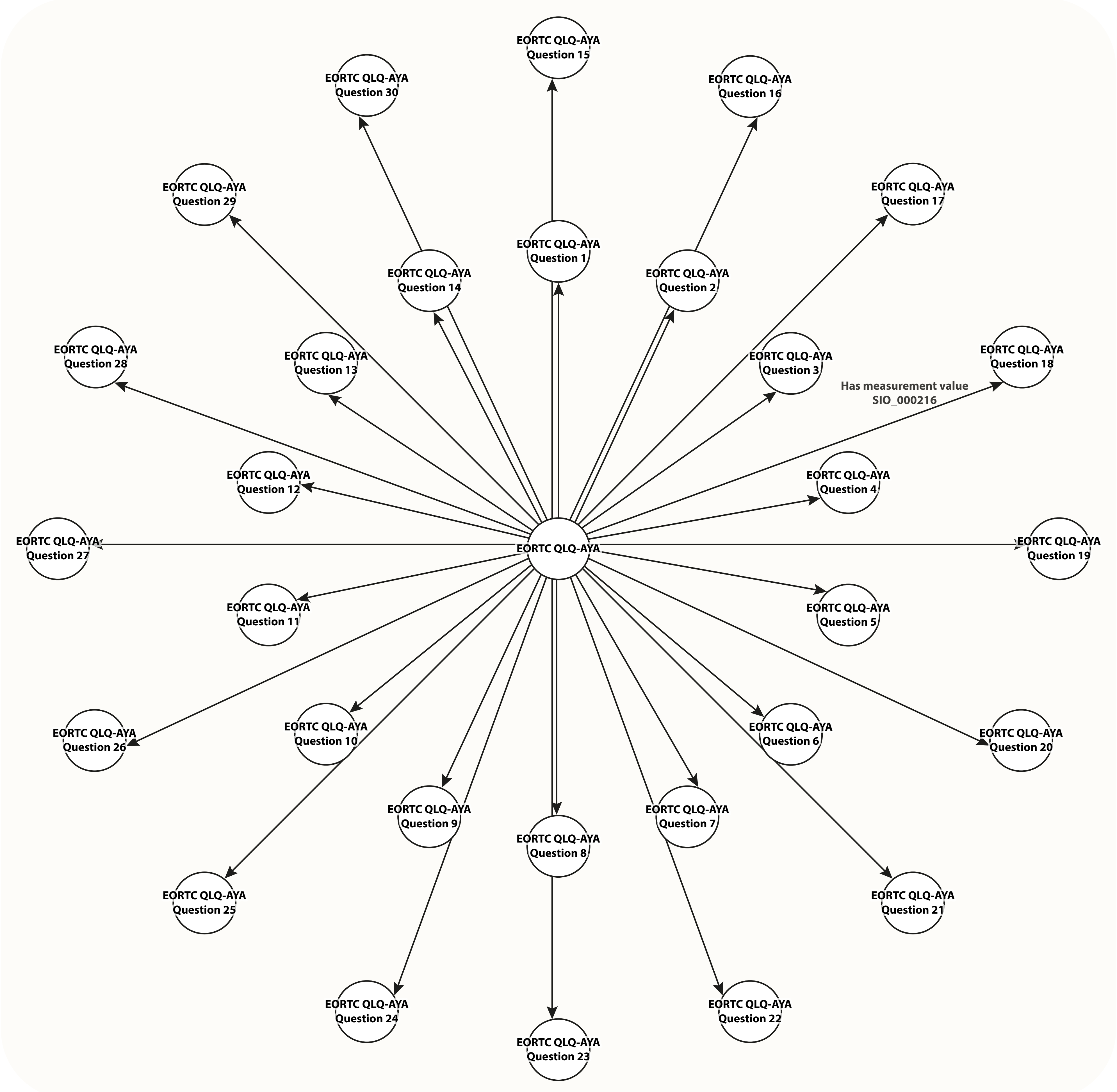


Supplementary Figure 2: Instrument graph illustrating the underlying measurements of a measurement instrument with multiple sub-concepts, such as here exemplified using the EORTC QLQ-AYA patient reported outcome measure.
Generally, instrument graphs comprise the main concept with its sub-concepts as measurement values– using SIO's 'has measurement value'.
EORTC QLQ-AYA refers to: European Organisation for Research and Treatment of Cancer Quality of Life Questionnaire Adolescent and Young Adults.
