## Supplementary Figure 3 for "Knowledge representation of a multi-centre adolescent and young adult (AYA) cancer infrastructure; development of the STRONG AYA Knowledge Graph"

**Mapping excerpt biological sex**

```
…
"biological_sex": {
 "predicate": "sio:SIO_000008",
 "class": "ncit:C28421",
 "local_definition": "alg_v1b",
 "schema_reconstruction": [
  {
   "type": "class",
   "predicate": "sio:SIO_000235",
   "class": "ncit:C18772",
   "class_label": "medicalClass",
   "aesthetic_label":
          "Medical_characteristics"
  },
  {
   "type": "class",
   "placement":"after",
   "predicate": "sio:SIO_000253",
   "class": "ncit:C95401",
   "class_label": "promClass",
   "aesthetic_label":
          "PROM"
  }
 ],
 "value_mapping": {
  "terms": {
   "male": {
    "local_term": "0.0",
    "target_class": "ncit:C20197"
   },
   "female": {
    "local_term": "1.0",
    "target_class": "ncit:C16576"
   },
   "intersex": {
    "local_term": null,
    "target_class": "ncit:C45908"
   },
   "missing_or_unspecified": {
    "local_term": null,
    "target_class": "ncit:C54031"
   }
  }
 }
},
…
```

Supplementary Figure 3: Mapping excerpt for synthetic SURVAYA biological sex, with local terminology in bold font.
