## Supplementary Figure 4 for "Knowledge representation of a multi-centre adolescent and young adult (AYA) cancer infrastructure; development of the STRONG AYA Knowledge Graph"

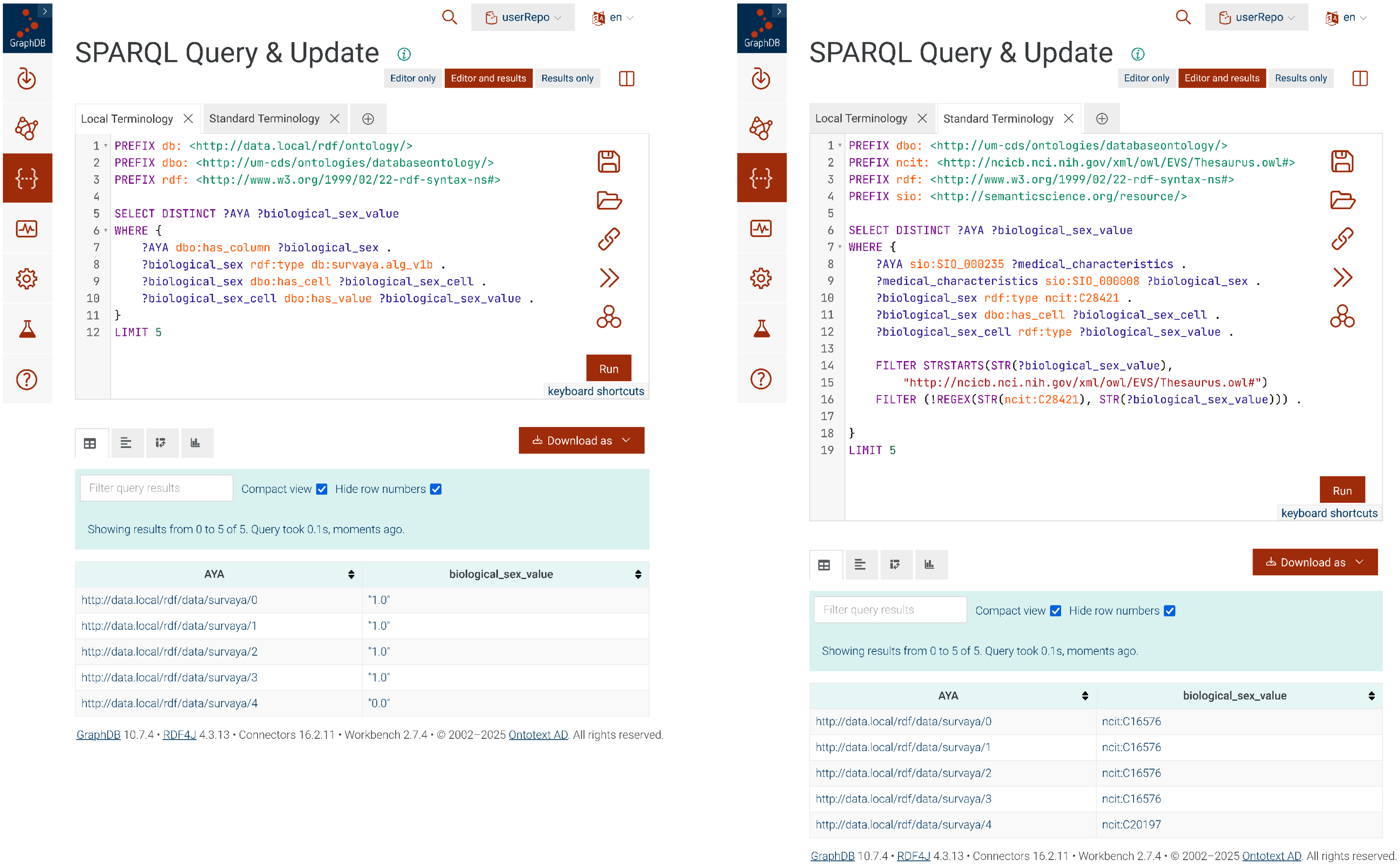


Supplementary Figure 4: Graph database output demonstrating the interoperability of our synthetic SURVAYA data.
